# Anthropometric measures and their relationship to corneal refractive power in the United States population

**DOI:** 10.1101/2021.09.10.21263400

**Authors:** Girish Valluru, Daniel Henick, Janek Klawe, Bian Liu, Louis R. Pasquale, Sumayya Ahmad

## Abstract

**Purpose:** To determine the relationship between anthropometric measures and corneal refractive power (CRP).

**Methods:** Participants from the 1999-2008 United States National Health and Nutrition Examination Survey (NHANES) visual exam with demographic, ocular, and anthropometric data (20,165 subjects) were included. Cases with steep cornea were defined by corneal power ≥ 48.0 diopters (n = 171) while controls had dioptric power < 48.0 D (n = 19,994). Multivariable analyses were performed for pooled and sex-stratified populations. Separate models assessed body mass index, height, and weight in relation to steep cornea.

**Results:** A relationship between BMI and steep cornea in the pooled population was not detected (P for trend = 0.78). There was a strong inverse relationship between height and steep cornea in the pooled population (P for trend <0.0001) and women (P for trend <0.0001). For every 1-inch increase in height, there was a 16% reduced odds of steep cornea in the pooled population (OR, 0.84; 95% CI: 0.78-0.91). There was also a significant inverse relationship between weight and steep cornea in the pooled population (P for trend = 0.01) and in men (P for trend = 0.02). For each 10-pound increase in weight there was a 7% reduced odds of steep cornea (OR, 0.927; 95% CI: 0.882-0.975) in the pooled analysis.

**Conclusions:** Greater height and greater weight were associated with a lower risk of steep cornea. These findings can contribute to an improved understanding of the pathogenesis of corneal ectasias.

## Introduction

Corneal refractive power (CRP) is an important clinical parameter in clinical practice for patients seeking refractive or cataract surgery. High corneal refractive power can indicate pathologic processes such as corneal ectasias like keratoconus and pellucid marginal degeneration.^1,2^ There is an intimate relationship between CRP and other ocular components such as axial length, anterior chamber depth, and lens thickness, with evidence that the eye often achieves emmetropia despite differences in these parameters.^3,4^ While there is a defined relationship between other ocular components and CRP, the relationship between CRP and anthropometric features is less clear.

Previous studies have explored these relationships and found variable results.^5–10^ Lee found an inverse relationship between corneal steepness and height after adjusting for confounders.^9^ Wong et al.’s population-based study in Singapore demonstrated the same results, as well as a positive relationship between weight and hyperopic refractions.^10^ While these studies suggest evidence of an inverse relationship between CRP/corneal radius and height, few papers have specifically examined anthropometric parameters in patients on patients with high CRP, a population which may represent those with corneal pathology. To date, the Central India Eye and Medical Study, the Beijing Eye study, and the Singapore Epidemiology of Eye Diseases Study have specifically examined anthropometric parameters on patients with steep corneas.^6–8^ However, these studies did not account for the confounding effects between the various anthropometric measures, had smaller sample sizes, and looked at ethnically homogenous populations.

The purpose of the current study was to investigate the relationship between anthropometric features and steep corneas (those with a spherical dioptric power of ≥ 48 D) using the National Health and Nutrition Examination Survey (NHANES). In doing so, we aimed to use methodology designed to assess the independent associations of height and of weight with CRP.

## Methods

### Data Sources

We used data from the 1999-2008 National Health and Nutrition Examination Survey (NHANES). The NHANES has been described previously. Briefly, it is a nationally representative survey of the US civilian, noninstitutionalized population with data on demographics, lifestyle, anthropometric measures, and also ocular health.^11^

The 1999-2008 NHANES was reviewed and approved by the National Center for Health Statistics Research Ethics Review Board. All participants provided written informed consent.

### Study Population Selection

We included all NHANES participants who met the following criteria: they took part in the visual exam; provided demographic data (age, gender, race, education, socioeconomic status) and general health information (on asthma, hypertension, and diabetes); anthropometric measures (height, weight); were older than 20 years (to account for continuing changes in height^12^); and had not had cataract or refractive eye surgery.

### Anthropometric Measurements

During visits at the mobile examination center, trained health technicians directly measured anthropometric features. A stadiometer was used to measure standing height to the nearest 0.1 centimeter. Weight was measured to the nearest 0.1 kilogram using a Toledo digital scale (Mettler-Toledo Inc.).

### Outcome Measures

Participants were examined using an autorefractor (NIDEK ARK-760; Nidek Co Ltd, Tokyo, Japan) in order to measure keratometry power in both eyes. For each eye, the dioptric power of the cornea was averaged across the meridians. Patients were examined in a non-cycloplegic state with final values from an average of three measurements.^13^ We defined steep cornea as a spherical dioptric power ≥ 48.0 diopters D in the eye with higher dioptric power. This cutoff was chosen for consistency with prior studies defining steep corneas as keratometry values ≥ 48 D.^6,8^

### Statistical Analysis

We used multivariable logistic regression models to assess the association of the primary exposure of an anthropometric measure (while adjusting for covariates of demographic factors and comorbidities as discussed further below) with the outcome variable of the presence of steep cornea. All analyses took into account NHANES survey design. SAS statistical software version 9.2 (SAS Institute, Cary, NC) and R statistical software version 1.1 (R Foundation, Vienna, Austria) were used for analysis.

### Anthropometric Measures

#### Uncorrelated Height and Weight analysis

Although BMI incorporates both height and weight, we were interested in assessing the independent associations of height and weight with steep corneas. Thus we performed a regression of height with weight as the exposure. This regression was used to calculate residuals of height that were centered at the median height of the population (66 inches) providing an uncorrelated height. For gender-specific analyses we repeated this process and centered at the median heights for men (69 inches) and women (64 inches). The resulting uncorrelated height and weight were included in a multivariable model to determine their association with steep cornea. This same process was repeated for weight residuals centered at the median weight of the entire population (172 lbs) and in gender-stratified analyses for men (187 lbs) and women (157 lbs).

### Regression Models

We fit multivariable logistic regression models with primary exposures of (1) BMI, (2) height, and (3) weight, and an outcome of the presence (vs. absence) of steep cornea. The first model had a primary exposure of BMI (bucketed into 5 categories), adjusting for gender, race, educational level, socioeconomic status, age, and comorbidities (asthma, HTN, and diabetes). The second model had a primary exposure of height adjusting for uncorrelated weight and the same covariates. The third model had a primary exposure of weight adjusting for uncorrelated height and the same covariates. These models were repeated with primary exposures as continuous variables to assess for linear relationships. Given the substantially larger range of weight as compared to height or BMI, weight was assessed as a continuous variable per 10 pounds. Outliers (1 case and 82 controls) in terms of measured data for BMI, height, and weight that were 3 times the interquartile range greater than the third quartile or lesser than the first quartile were not included in the analysis shown.^14^ Sensitivity analysis was conducted with outliers included (data not shown) and is further described in the results.

In order to assess the role of gender with each of the three primary exposures, three approaches were used: (1) gender was adjusted for as a covariate in multivariable regressions with the pooled analyses; (2) multivariable models with an interaction term between gender and the primary exposure were assessed; and (3) models were assessed for the entire population and then repeated with gender stratification.

## Results

Among 20,165 individuals, 171 cases of steep corneas were identified. In the overall population (Table 1), the mean age for controls was 46 years (range, 20-85; 95% CI 45.8-46.9) and for cases was 44 years (range, 20-85; 95% CI 41.4-47.1). Women comprised 52% (95% CI 50.9-52.2) of the overall population. Notably, there were more women among cases (68%; 95% CI 59.2-76.8) than controls (51%;95% CI 50.8-52.0). There were also more individuals with asthma in cases (23%; 95% CI 14.0-32.5) than among controls (13%; 95% CI 12.2-13.5).

**Table 1:** Demographic and other characteristics of the National Health and Nutrition Examination Survey sample stratified by keratometry readings

| Characteristics | Spherical power < 48 diopters |  | Spherical power ≥ 48.0 diopters |  |
| --- | --- | --- | --- | --- |
|  | Number | Percentage | Number | Percentage |
| **Gender** |  |  |  |  |
| Male | 9687 | 48.6 | 58 | 32.0 |
| Female | 10307 | 51.4 | 113 | 68.0 |
| **Age (years)** |  |  |  |  |
| 20 - 39 | 7247 | 39.3 | 63 | 41.2 |
| 40 - 59 | 6166 | 39.1 | 53 | 38.1 |
| ≥ 60 | 6581 | 21.6 | 55 | 20.7 |
| **Race** |  |  |  |  |
| Mexican American | 4002 | 7.2 | 34 | 6.7 |
| Other Hispanic | 1098 | 4.8 | 8 | 2.8 |
| Non-Hispanic white | 10229 | 72.7 | 97 | 77.2 |
| Non-Hispanic black | 3930 | 10.4 | 29 | 10.5 |
| Other race | 735 | 4.8 | 3 | 2.9 |
| **Education** |  |  |  |  |
| Less than High School | 5821 | 18.5 | 49 | 20.3 |
| High School | 4811 | 25.5 | 48 | 26.8 |
| Greater than High School | 9362 | 56.1 | 74 | 52.9 |
| **Socioeconomic Status\*** |  |  |  |  |
| Above Poverty Line | 16341 | 87.1 | 131 | 83.8 |
| Below Poverty Line | 3653 | 12.9 | 40 | 16.2 |
| **Comorbidities\*** |  |  |  |  |
| Asthma | 2399 | 12.9 | 32 | 23.3 |
| High blood pressure | 6393 | 27.8 | 55 | 25.9 |
| Diabetes | 2041 | 7.1 | 21 | 9.3 |
| **Height (inches)** |  |  |  |  |
| ≤ 62 | 3260 | 12.2 | 59 | 27.3 |
| 62.1-65 | 5075 | 24.4 | 48 | 31.4 |
| 65.1-67 | 3643 | 18.1 | 25 | 15.7 |
| 67.1-70 | 4610 | 24.3 | 21 | 12.6 |
| 70.1+ | 3406 | 21.1 | 18 | 13.1 |
| **Weight (pounds)** |  |  |  |  |
| ≤ 140 | 3937 | 19.7 | 50 | 30.3 |
| 140.1-160 | 3756 | 17.7 | 33 | 18.5 |
| 160.1-180 | 4004 | 19.3 | 32 | 17.2 |
| 180.1-200 | 3205 | 16.0 | 24 | 12.7 |
| 200.1 + | 5092 | 27.3 | 32 | 21.3 |
| **Body Mass Index (kg/m<sup>2</sup>)** |  |  |  |  |
| ≤ 22 | 2427 | 13.8 | 23 | 14.7 |
| 22.1-25 | 3772 | 20.1 | 30 | 19.4 |
| 25.1-27 | 3018 | 14.8 | 23 | 13.6 |
| 27.1-29 | 2804 | 13.3 | 24 | 15.1 |
| 29.1 + | 7973 | 38.1 | 71 | 37.3 |

\*Socioeconomic status is calculated by the Center for Disease Control as a ratio of family income to the poverty line based on annual poverty guidelines specific to family size and geographic location issued by the Department of Health and Human services. Comorbidities are self-reported by patients based on if a physician has ever told them they have the condition (i.e. asthma, hypertension, diabetes).

There were notable differences in sex-stratified analysis (Table S1). Among women, there were more cases with asthma (28%; 95% CI 16.6-39.2) than controls (15%; 95% CI 13.6-15.5). Among men, there were more cases who were non-Hispanic whites (83%;95% CI 74.5-90.7) than controls (73%; 95% CI 70.2-75.3). All differences were adjusted for in multivariable analysis.

### The relationship between body-mass index and steep cornea (Table 2)

A relationship between categories of BMI and steep cornea was not detected in the pooled population in categorical analysis (p for trend = 0.78). A relationship between linear BMI and steep cornea was also not detected (p=0.86).

**Table 2.**
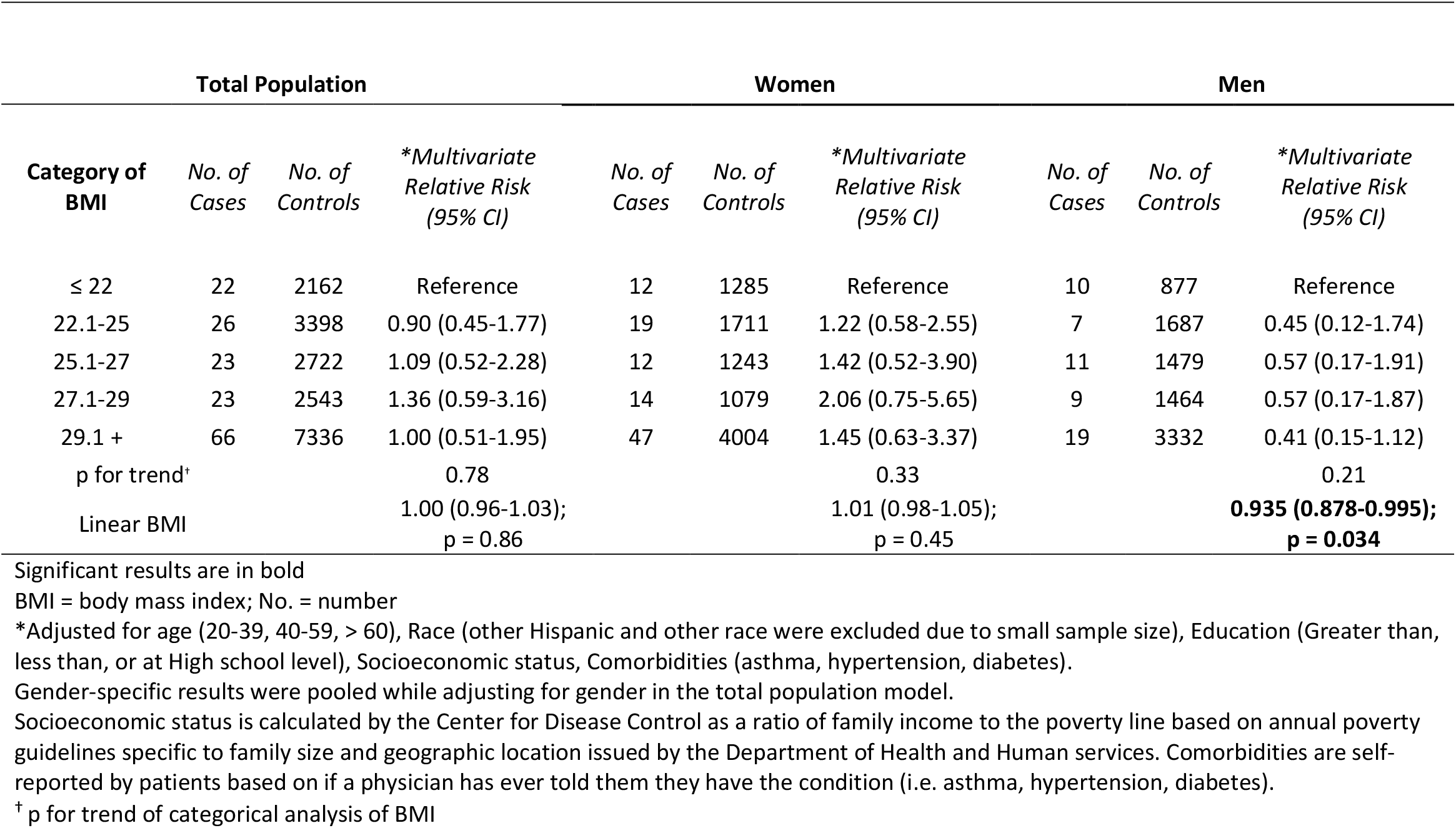
Pooled and gender specific analysis of body mass index (kg/m^2^) in relation to steep cornea defined by high spherical dioptric power (≥ 48 D)

|  |  |  |  |  |  |  |  |  |  |
| --- | --- | --- | --- | --- | --- | --- | --- | --- | --- |
| Total Population |  |  |  | Women |  |  | Men |  |  |
| Category of BMI | No. of Cases | No. of Controls | *Multivariate Relative Risk (95% CI) | No. of Cases | No. of Controls | *Multivariate Relative Risk (95% CI) | No. of Cases | No. of Controls | *Multivariate Relative Risk (95% CI) |
| ≤ 22 | 22 | 2162 | Reference | 12 | 1285 | Reference | 10 | 877 | Reference |
| 22.1-25 | 26 | 3398 | 0.90 (0.45-1.77) | 19 | 1711 | 1.22 (0.58-2.55) | 7 | 1687 | 0.45 (0.12-1.74) |
| 25.1-27 | 23 | 2722 | 1.09 (0.52-2.28) | 12 | 1243 | 1.42 (0.52-3.90) | 11 | 1479 | 0.57 (0.17-1.91) |
| 27.1-29 | 23 | 2543 | 1.36 (0.59-3.16) | 14 | 1079 | 2.06 (0.75-5.65) | 9 | 1464 | 0.57 (0.17-1.87) |
| 29.1 + | 66 | 7336 | 1.00 (0.51-1.95) | 47 | 4004 | 1.45 (0.63-3.37) | 19 | 3332 | 0.41 (0.15-1.12) |
| p for trend† |  |  | 0.78 |  |  | 0.33 |  |  | 0.21 |
| Linear BMI |  |  | 1.00 (0.96-1.03);<br>p = 0.86 |  |  | 1.01 (0.98-1.05);<br>p = 0.45 |  |  | **0.935 (0.878-0.995);**<br>**p = 0.034** |

Significant results are in bold
BMI = body mass index; No. = number
\*Adjusted for age (20-39, 40-59, > 60), Race (other Hispanic and other race were excluded due to small sample size), Education (Greater than, less than, or at High school level), Socioeconomic status, Comorbidities (asthma, hypertension, diabetes).
Gender-specific results were pooled while adjusting for gender in the total population model.
Socioeconomic status is calculated by the Center for Disease Control as a ratio of family income to the poverty line based on annual poverty guidelines specific to family size and geographic location issued by the Department of Health and Human services. Comorbidities are self-reported by patients based on if a physician has ever told them they have the condition (i.e. asthma, hypertension, diabetes).
<sup>†</sup> p for trend of categorical analysis of BMI

In sex stratified analysis, there was a nonsignificant negative relationship between categorical BMI and odds of steep cornea in men (p for trend = 0.21). When BMI was considered as a continuous variable, each unit increase in BMI was associated with a 6% reduced odds of steep cornea in men (OR, 0.94; 95% CI 0.878-0.995). While inclusion of outliers did change the borderline significance of this association, it did not meaningfully change the reduced odds of steep cornea in men per unit increase in BMI (OR, 0.95; 95% CI 0.889-1.006). Analysis of the BMI categories (p = 0.33, for trend) and of linear BMI showed no significant associations with steep cornea in women (p = 0.45). When using the existing categories (Table S1), there was not a significant interaction between BMI and gender in association with steep cornea (p = 0.41, Table S2) in categorical analysis.

### The relationship between height and steep cornea (Table 3)

There was an inverse relationship between categories of height (adjusted for weight residuals) and steep cornea (p for trend <0.0001) in the pooled population. Compared to participants ≤ 62 inches, for those in the 65.1 – 67-inch category there was a 61% reduced odds (OR, 0.29; 95% CI 0.16-0.55) of steep cornea and in the tallest category ≥ 70.1 inches there was a 82% reduced odds (OR, 0.18; 95% CI 0.07-0.47). The relation between linear height and steep cornea was robust (p<0.0001). For every 1-inch increase in height adjusted for weight residuals, there was a 16% reduced odds of steep cornea (OR, 0.84; 95% CI 0.78-0.91).

**Table 3.** Pooled and gender specific analysis of height adjusted for weight residuals in relation to steep cornea defined by high spherical dioptric power (≥ 48 D).

| Total Population |  |  |  | Women |  |  |  | Men |  |  |  |
| --- | --- | --- | --- | --- | --- | --- | --- | --- | --- | --- | --- |
| Category of Height (inches) | No. of Cases | No. of Controls | *Multivariate Relative Risk (95% CI) | Category of Height (inches) | No. of Cases | No. of Controls | *Multivariate Relative Risk (95% CI) | Category of Height (inches) | No. of Cases | No. of Controls | *Multivariate Relative Risk (95% CI) |
| $\leq 62$ | 53 | 2765 | Reference | $\leq 61$ | 31 | 1677 | Reference | $\leq 66$ | 15 | 1517 | Reference |
| 62.1-65 | 46 | 4580 | **0.50 (0.31-0.81)** | 61.1-63 | 34 | 2222 | 0.70 (0.36-1.39) | 66.1-68 | 9 | 1875 | 0.30 (0.09-1.03) |
| 65.1-67 | 22 | 3302 | **0.29 (0.16-0.55)** | 63.1-64 | 13 | 1292 | **0.46 (0.24-0.88)** | 68.1-69 | 10 | 1132 | 0.67 (0.20-2.16) |
| 67.1-70 | 21 | 4245 | **0.17 (0.08-0.36)** | 64.1-66 | 17 | 2376 | **0.35 (0.18-0.71)** | 69.1-71 | 8 | 2196 | **0.25 (0.09-0.70)** |
| 70.1+ | 18 | 3269 | **0.18 (0.07-0.47)** | 66.1+ | 9 | 1755 | **0.23 (0.10-0.55)** | 71.1 + | 14 | 2119 | 0.49 (0.14-1.73) |
| p for trend<sup>†</sup> |  |  | **< 0.0001** |  |  |  | **< 0.0001** |  |  |  | 0.46 |
| Linear Height |  |  | **0.84 (0.78-0.91);**<br>**p < 0.0001** |  |  |  | **0.81 (0.74-0.88);**<br>**p < 0.0001** |  |  |  | 0.88 (0.76-1.01);<br>p = 0.074 |

Significant results are in bold. No. = number
\*Adjusted for weight residuals (5 categories), age (20-39, 40-59, > 60 years), Race (other Hispanic and other race were excluded due to small sample size), Education (Greater than, less than, or at High school level), Socioeconomic status, Comorbidities (asthma, hypertension, diabetes).
Gender-specific results were pooled while adjusting for gender in the total population model.
Socioeconomic status is calculated by the Center for Disease Control as a ratio of family income to the poverty line based on annual poverty guidelines specific to family size and geographic location issued by the Department of Health and Human services. Comorbidities are self-reported by patients based on if a physician has ever told them they have the condition (i.e. asthma, hypertension, diabetes).
<sup>†</sup> p for trend of categorical analysis of height

In gender specific analysis, this relationship between height adjusted for weight residuals and steep cornea was also significant in women. For every 1-inch increase in height there was a 19% reduced odds of steep cornea (OR, 0.81; 95% CI 0.74-0.88; p<0.0001).

The relationship between height adjusted for weight residuals and steep cornea in men showed equivocal results. In multivariable analysis, only one category of height (69.1-71 inches) showed an inverse relationship with steep cornea (OR, 0.25; 95% CI: 0.09-0.70) compared to the reference category (≤66 inches) in men. There was a borderline significant association between height and steep cornea in men when assessed for a linear relationship (OR, 0.88; 95% CI 0.76-1.01; p =0.074).

We explored a potential gender-height interaction in relation to steep cornea in several multivariable models. When height was treated as a continuous variable, no significant interaction was found (interaction coefficient -0.03; 95% CI -0.08 to 0.03, p=0.35; Table S3). Further, when we readjusted the categories for the inherent differences in height between men and women no significant interaction was noted (p= 0.59; Table S5).

### The relationship between weight and steep cornea (Table 4)

There was an inverse relationship between categories of weight (adjusted for height residuals) and steep cornea (p for trend = 0.011) in the pooled population. While there is a trend between categories of weight and their association with steep cornea, only the highest category of weight (200.1 lbs+) showed a statistically significant inverse relationship with steep cornea (OR, 0.45; 95% CI: 0.24-0.85) compared to the reference category (≤140 lbs). When weight was considered as a linear variable, every 10-pound increase in weight had a 7% reduced odds of steep cornea (OR, 0.927; 95% CI: 0.882-0.975; p= 0.0038).

**Table 4.** Pooled and gender-specific analysis of weight adjusted for height residuals in relation to steep cornea defined by high spherical dioptric power (≥ 48 D)

| Total Population |  |  |  | Women |  |  |  | Men |  |  |  |
| --- | --- | --- | --- | --- | --- | --- | --- | --- | --- | --- | --- |
| Category of weight (pounds) | No. of Cases | No. of Controls | *Multivariate Relative Risk (95% CI) | Category of weight (pounds) | No. of Cases | No. of Controls | *Multivariate Relative Risk (95% CI) | Category of weight (pounds) | No. of Cases | No. of Controls | *Multivariate Relative Risk (95% CI) |
| $\leq 140$ | 45 | 3401 | Reference | $\leq 128$ | 23 | 1552 | Reference | $\leq 153$ | 12 | 1400 | Reference |
| 140.1 - 160 | 31 | 3371 | 0.73 (0.41-1.31) | 128.1-148 | 20 | 1981 | 0.73 (0.34-1.55) | 153.1-173 | 11 | 1774 | 0.64 (0.20-2.09) |
| 160.1 - 180 | 29 | 3657 | 0.61 (0.30-1.24) | 148.1-168 | 24 | 1975 | 0.94 (0.45-1.94) | 173.1-182 | 11 | 969 | 1.28 (0.38-4.29) |
| 180.1 - 200 | 24 | 2966 | 0.56 (0.31-1.01) | 168.1-188 | 12 | 1488 | 0.56 (0.23-1.38) | 182.1-202 | 10 | 1798 | 0.43 (0.16-1.18) |
| 200.1 + | 31 | 4766 | **0.45 (0.24-0.85)** | 188.1 + | 25 | 2326 | 0.90 (0.47-1.74) | 202.1 + | 12 | 2898 | 0.38 (0.13-1.08) |
| p for trend<sup>†</sup> |  |  | **p = 0.011** |  |  |  | p = 0.75 |  |  |  | **p = 0.024** |
| Linear weight (per pound) |  |  | **0.992 (0.987-0.997)** |  |  |  | 0.999 (0.992-1.005) |  |  |  | **0.988 (0.979-0.997)** |
| Linear weight (per 10 pounds) |  |  | **0.927 (0.882-0.975);**<br>**p = 0.0038** |  |  |  | 0.986 (0.926-1.049);<br>p = 0.64 |  |  |  | **0.885 (0.809-0.968);**<br>**p = 0.0083** |

Significant results are in bold. No. = number
\*Adjusted for height residuals (5 categories), age (20-39, 40-59, > 60 years), Race (other Hispanic and other race were excluded due to small sample size), Education (Greater than, less than, or at High school level), Socioeconomic status, Comorbidities (asthma, hypertension, diabetes).
Gender-specific results were pooled while adjusting for gender in the total population model.
Socioeconomic status is calculated by the Center for Disease Control as a ratio of family income to the poverty line based on annual poverty guidelines specific to family size and geographic location issued by the Department of Health and Human services. Comorbidities are self-reported by patients based on if a physician has ever told them they have the condition (i.e. asthma, hypertension, diabetes).
<sup>†</sup> p for trend of categorical analysis of weight

In sex stratified analysis, there was not a significant association between weight adjusted for height residuals and steep cornea in women in both categorical (p= 0.75) and linear analyses per 10-pound increase (p= 0.64). Among men, for each 10-pound increase in weight there was 12% reduced odds of steep cornea (OR, 0.885; 95% CI: 0.809-0.968; p=0.0083). While there were modest differences in the associations of steep cornea and weight in sex-stratified analysis, there was not a significant interaction between weight and gender in relationship to steep cornea (interaction coefficient 0.02; 95% CI -0.01 to 0.04, p = 0.16, Table S4).

## Discussion

This retrospective cross-sectional study found a substantial inverse association between height and steep cornea. Notably, this relationship, when adjusting for uncorrelated weight, was significant in the total population and in women. Weight, adjusted for uncorrelated height, was also inversely associated with steep cornea in the total population and in a sex-stratified analysis for men, but not for women. There was no significant association between BMI and steep cornea.

Height had the strongest association with steep cornea; for each 1-inch increase in height there were 16% lower odds of steep cornea in the pooled population, while in women there was a 19% reduction. Studies in India and Singapore have identified a similar inverse relationship between height and corneas with K>48, even after adjusting for other variables.^6–8^ The Beiijng eye study had similar results until adjustment for other ocular characteristics (i.e. axial length, cylindrical refractive error, etc.). None of these three studies used residuals to de-correlate the height and weight variables. Our study, in context of those that have shown a similar relationship, suggest a possible genetic relationship between height and steep corneas.

There is a complex connection between corneal curvature/steepness and other ocular features such as axial length and anterior chamber depth. Longer axial lengths are often associated with flatter corneas and shorter axial lengths are associated with steeper corneas, in an effort of the eye to achieve emmetropization. It has been theorized that those with shorter height may also have smaller axial lengths, and secondarily develop steeper corneas. It has also been speculated that educational status is associated within better nutrition and taller height, which may lead to myopia and longer axial length. Previous researchers have examined these relationships.^7,9,10^ Wong et al. found that that height had a strong correlation to multiple ocular parameters (axial length, corneal curvature, anterior chamber and vitreous chamber depth) that is independent of education, weight, gender, and other systemic and lifestyle factors. Lee et al also found that height was independently associated with corneal curvature after controlling for education, gender, and age.^9^ Our study also showed that the effect of height persists after adjusting for education, race, and socioeconomic status, challenging the hypothesis of education being a surrogate marker for height, better nutrition and longer axial lengths.

Although other studies used the term “keratoconus (KCN)” interchangeably with K >48, we felt that using this definition would mis-classify many individuals. Keratoconus is a clinical diagnosis that relies on both the clinical exam and corneal imaging findings (either tomography or topography). Using a diopter cut-off value may result in people with corneal scars being incorrectly labeled as having KCN, but also may miss those with early disease. Nonetheless, many people in our analysis likely did have true KCN, and the relationship between KCN and height warrants discussion. There are known associations between connective tissue disorders, with varying height relationships, and KCN, suggesting an underlying role of connective tissue dysfunction.^2,15^ Notably, reports of greater prevalence of mitral valve prolapse in individuals with KCN when compared to controls suggest that alterations in collagen subtypes during embryogenesis may lead to changes in the corneal stroma as well as mitral valve formation.^15–17^ There may be a genetic basis for the association between height and KCN as both height and central corneal thickness (strongly associated with KCN) have substantial heritability with estimates up to 80% and 95%, respectively.^18,19^ Further study of this association may elucidate these genetic underpinnings.

Estrogen may contribute to the association of steeper corneas and shorter height. Estrogen plays an important role in the development of height promoting early epiphyseal growth plate closure and resulting in shorter height.^20,21^ In genetic studies, estrogen receptor polymorphisms have been associated with decreased height.^22,23^ Healthy women have also demonstrated changes in corneal curvature and thickness during the menstrual cycle, menopause, and with topical estrogen therapy.^24–26^ Although most studies on estrogen and corneal topographic changes are predominantly in women, further study is needed to characterize estrogen effects on corneal topography in men.

While our analyses did not identify notable interactions between anthropometric features and gender in relationship to steep cornea, this may have been due to limited sample size and further in the case of height, inherent gender differences in distributions. Prior studies have reported the relationship between gender and steep cornea and anthropometric features.^6–8^ Some studies found that once height was included, a gender association with steep cornea was no longer significant, similar to what we found.^6–8^ Our findings on the confounding effects of height or weight on the relationship between gender and steep cornea (see supplement) may provide insight on these differing associations.^27–29^ Further investigation with case-control studies may enable detection of associations and interactions between anthropometric features and gender in people with steep corneas.

In addition to the inverse relationship between height and steep cornea, we also found a significant association between weight and steep cornea; there was a 7% reduced odds of steep cornea per 10 pound increase in weight. Similarly, in men there was a 12% reduced odds of steep cornea per 10 pound increase in weight. A potential biological basis for this relationship may be related to hormones. Prolactin, which is found at higher levels in patients with obesity, and the related prolactin induced protein, may mediate transcription, translation, and organization of keratin in the cornea.^30–33^ Interestingly, prolactin induced protein levels are downregulated in tears, saliva, and corneal fibroblast cells from individuals with KCN compared to healthy individuals.^30^

There was no meaningful association between BMI and steep cornea in our pooled analysis. In men, a modest association between BMI and steep cornea was detected; they were found to have a 6% reduced odds of steep cornea per unit increase in BMI. Finally, there were no significant categorical interactions with gender and BMI (p = 0.41) or gender and weight (p = 0.61) in steep cornea. There are mixed findings in the literature regarding the relationship between steep cornea and BMI.^6–8^ Although some studies^6–8^ similarly did not find an association between BMI and steep cornea, another paper from Israel by Eliasi et al. found a strong relationship between KCN (as defined by patients with K >48) and BMI. ^5^ While both our study and that of Eliasi adjust for age, their narrow age inclusion criteria (16-19.9 years old) may be contributing to the differences in our findings.

Our study has several limitations. First, although we adjusted our models for a number of important confounding factors, the NHANES did not collect other ocular parameters such as CCT or axial length, which have been included as covariates in other studies.^6–8^ This may be needed to better understand their role in this association. Finally, we cannot draw causal inferences from our analyses with the cross-sectional data of the NHANES. We acknowledge this limitation and further studies using different methodologies will be needed to address causal relationships.

Strengths of this study included the use of uncorrelated height and weight in analyses to assess their independent associations with steep cornea. This allowed greater power to detect associations with steep cornea and enables stronger associations to be identified. The NHANES has highly standardized and calibrated methodology allowing for accurate and direct measurements of anthropometric features and keratometry, both of which were measured in the same visit. ^34^ Further, our analysis used multiple logistic regression to account for a number of covariates including age, race, education, socioeconomic status, and comorbidities (asthma, hypertension, and diabetes). Finally, the NHANES database multilevel complex sampling methodology enables findings applicable to a nationally representative US population.

In conclusion, there is an inverse association between height, independent of weight, and steep cornea. There is also an inverse relationship between weight, independent of height, and steep cornea. Further study of the relationships of height and weight with steep cornea may yield mechanistic insight into the pathogenesis of corneal ectasias as well as genetic correlations.

## Data Availability

We used data from the 1999-2008 National Health and Nutrition Examination Survey (NHANES). The NHANES has been described previously. Briefly, it is a nationally representative survey of the US civilian, noninstitutionalized population with data on demographics, lifestyle, anthropometric measures, and also ocular health (see citation below).
Engel A, Murphy RS, Maurer K, Collins E. Plan and operation of the HANES I augmentation survey of adults 25-74 years United States, 1974-1975 - PubMed. Accessed December 19, 2020. https://pubmed-ncbi-nlm-nih-gov.eresources.mssm.edu/695333/

## Supplemental Tables

As described in methods on regression models, analysis of interactions between gender and anthropometric features was conducted. An interaction term between gender and an anthropometric feature (Table S2-S4) was included in regression models. Height categories were additionally adjusted for inherent differences in height between men and women (Table S5). Quintiles by height were calculated separately for women and then men in order to adjust for inherent gender differences in height. 28

**Table S1:** Gender specific demographic and other characteristics of the National Health and Nutrition Examination Survey sample stratified by keratometry readings

| Characteristics | Men |  |  |  | Women |  |  |  |
| --- | --- | --- | --- | --- | --- | --- | --- | --- |
|  | Spherical power<br>< 48 diopters |  | Spherical power<br>≥ 48.0 diopters |  | Spherical power<br>< 48 diopters |  | Spherical power<br>≥ 48.0 diopters |  |
|  | Number | Percentage | Number | Percentage | Number | Percentage | Number | Percentage |
| **Age (years)** |  |  |  |  |  |  |  |  |
| 20 – 39 | 3280 | 40.4 | 17 | 41.0 | 3967 | 38.2 | 46 | 41.3 |
| 40 – 59 | 3098 | 39.8 | 16 | 36.0 | 3068 | 38.5 | 37 | 39.0 |
| ≥ 60 | 3309 | 19.8 | 25 | 23.0 | 3272 | 23.3 | 30 | 19.6 |
| **Race** |  |  |  |  |  |  |  |  |
| Mexican American | 1920 | 8.0 | 13 | 8.1 | 2082 | 6.5 | 21 | 6.0 |
| Other Hispanic | 505 | 4.7 | 2 | 1.7 | 593 | 5.0 | 6 | 3.2 |
| Non-Hispanic white | 5021 | 72.8 | 36 | 82.6 | 5208 | 72.6 | 61 | 74.7 |
| Non-Hispanic black | 1898 | 9.7 | 7 | 7.6 | 2032 | 11.1 | 22 | 11.9 |
| Other race | 343 | 4.8 | 0 | 0 | 392 | 4.8 | 3 | 4.2 |
| **Education** |  |  |  |  |  |  |  |  |
| Less than High School | 2957 | 19.2 | 17 | 14.2 | 2864 | 17.8 | 32 | 23.1 |
| High School | 2337 | 25.8 | 19 | 38.1 | 2474 | 25.2 | 29 | 21.5 |
| Greater than High School | 4393 | 55.0 | 22 | 47.6 | 4969 | 57.0 | 52 | 55.4 |
| **Socioeconomic Status\*** |  |  |  |  |  |  |  |  |
| Above Poverty Line | 8077 | 88.8 | 42 | 81.6 | 8264 | 85.5 | 89 | 84.9 |
| Below Poverty Line | 1610 | 11.2 | 16 | 18.4 | 2043 | 14.5 | 24 | 15.1 |
| **Comorbidities\*** |  |  |  |  |  |  |  |  |
| Asthma | 992 | 11.1 | 6 | 13.4 | 1407 | 14.6 | 26 | 27.9 |
| High blood pressure | 3041 | 26.5 | 22 | 29.4 | 3352 | 29.0 | 33 | 24.2 |
| Diabetes | 1037 | 7.2 | 10 | 9.9 | 1004 | 7.1 | 11 | 9.0 |
| **Height (inches)** |  |  |  |  |  |  |  |  |
| ≤ 62 | 117 | 0.7 | 3 | 3.4 | 3143 | 23.0 | 56 | 38.5 |
| 62.1-65 | 949 | 6.0 | 7 | 11.4 | 4126 | 41.7 | 41 | 40.8 |
| 65.1-67 | 1635 | 13.2 | 13 | 18.0 | 2008 | 22.7 | 12 | 14.6 |
| 67.1-70 | 3676 | 38.0 | 17 | 26.3 | 934 | 11.3 | 4 | 6.1 |
| 70.1 + | 3310 | 42.2 | 18 | 40.9 | 96 | 1.2 | 0 | 0 |
| **Weight (pounds)** |  |  |  |  |  |  |  |  |
| ≤ 140 | 774 | 6.4 | 7 | 32.4 | 3163 | 13.2 | 43 | 38.4 |
| 140.1-160 | 1507 | 13.7 | 12 | 21.5 | 2249 | 16.9 | 21 | 19.3 |
| 160.1-180 | 2148 | 21.4 | 14 | 17.3 | 1856 | 28.4 | 18 | 11.9 |
| 180.1-200 | 2022 | 21.3 | 11 | 10.9 | 1183 | 15.5 | 13 | 11.4 |
| 200.1 + | 3236 | 37.2 | 14 | 18.0 | 1856 | 26.0 | 18 | 19.0 |
| **Body Mass Index (kg/m2)** |  |  |  |  |  |  |  |  |
| ≤ 22 | 978 | 9.9 | 10 | 17.6 | 1449 | 17.4 | 13 | 13.3 |
| 22.1-25 | 1871 | 19.6 | 7 | 15.7 | 1901 | 20.6 | 23 | 21.2 |
| 25.1-27 | 1632 | 16.8 | 11 | 17.3 | 1386 | 12.9 | 12 | 11.9 |
| 27.1-29 | 1596 | 16.1 | 9 | 17.1 | 1208 | 10.7 | 15 | 14.1 |
| 29.1 + | 3610 | 37.6 | 21 | 32.3 | 4363 | 38.5 | 50 | 39.6 |

\*Socioeconomic status is calculated by the Center for Disease Control as a ratio of family income to the poverty line based on annual poverty guidelines specific to family size and geographic location issued by the Department of Health and Human services. Comorbidities are self-reported by patients based on if a physician has ever told them they have the condition (i.e. asthma, hypertension, diabetes).

**Table S2.** Analysis of Interaction between Gender and Body mass index (kg/m2) in relation to steep cornea defined by high spherical dioptric power (> 48 D) and normal level of spherical dioptric power (≤ 48 D)

| Category of BMI (kg/m<sup>2</sup>) | Men |  | Women |  | Categorical Interaction* |  |
| --- | --- | --- | --- | --- | --- | --- |
|  | No. of Cases | No. of Controls | No. of Cases | No. of Controls | Interaction coefficient (95% CI) | p-value |
| ≤ 22 | 10 | 877 | 12 | 1285 | Reference |  |
| 22.1-25 | 7 | 1687 | 19 | 1711 | 0.96 (-0.59, 2.51) | 0.23 |
| 25.1-27 | 11 | 1479 | 12 | 1243 | 0.84 (-0.86, 2.55) | 0.33 |
| 27.1-29 | 9 | 1464 | 14 | 1079 | 1.15 (-0.21, 2.52) | 0.10 |
| 29.1+ | 19 | 3332 | 47 | 4004 | 1.12 (-0.13, 2.37) | 0.08 |
| p for categorical interaction |  |  |  |  |  | 0.41 |
| Linear BMI |  |  |  |  | **0.07 (0.01, 0.13)** | **0.035** |

BMI = body mass index. No. = number
\*Adjusted for gender (men were the reference group), age (20-39, 40-59, > 60), Race (other Hispanic and other race were excluded due to small sample size), Education (Greater than, less than, or at High school level), Socioeconomic status, Comorbidities (asthma, hypertension, diabetes).
Socioeconomic status is calculated by the Center for Disease Control as a ratio of family income to the poverty line based on annual poverty guidelines specific to family size and geographic location issued by the Department of Health and Human services. Comorbidities are self-reported by patients based on if a physician has ever told them they have the condition (i.e. asthma, hypertension, diabetes).

**Table S3.** Analysis of Interaction between Gender and Height (inches) in relation to steep cornea defined by high spherical dioptric power (> 48 D) and normal level of spherical dioptric power (≤ 48 D)

| Table S3. Analysis of Interaction between Gender and Height (inches) in relation to steep cornea defined by high spherical dioptric power (> 48 D) and normal level of spherical dioptric power ($\leq 48$ D) | | | | |
| --- | --- | --- | --- | --- |
| Category of Height (inches) | Men |  | Women |  |
|  | No. of Cases | No. of Controls | No. of Cases | No. of Controls |
| <62 | 3 | 90 | 50 | 2675 |
| 62.1-65 | 7 | 797 | 39 | 3783 |
| 65.1-67 | 11 | 1414 | 11 | 1888 |
| 67.1-70 | 17 | 3363 | 4 | 882 |
| 70.1+ | 18 | 3175 | 0 | 94 |
|  |  |  | Interaction coefficient (95% CI)* | p-value |
| Linear height |  |  | -0.03 (-0.08, 0.03) | 0.35 |

No. = number
\*Adjusted for gender (men were the reference group), weight residuals, age (20-39, 40-59, > 60), Race (other Hispanic and other race were excluded due to small sample size), Education (Greater than, less than, or at High school level), Socioeconomic status, Comorbidities (asthma, hypertension, diabetes).
Socioeconomic status is calculated by the Center for Disease Control as a ratio of family income to the poverty line based on annual poverty guidelines specific to family size and geographic location issued by the Department of Health and Human services. Comorbidities are self-reported by patients based on if a physician has ever told them they have the condition (i.e. asthma, hypertension, diabetes).
Non-overlapping height distributions in men and women limited categorical interaction analysis. We sought to instead use continuous height to assess this interaction. Additional analysis adjusting for inherent gender differences is shown in table S5.

**Table S4.** Analysis of Interaction between Gender and Weight (pounds) in relation to steep cornea defined by high spherical dioptric power (> 48 D) and normal level of spherical dioptric power (≤ 48 D)

| Category of Weight (pounds) | Men |  | Women |  | Categorical Interaction* |  |
| --- | --- | --- | --- | --- | --- | --- |
|  | No. of Cases | No. of Controls | No. of Cases | No. of Controls | Interaction coefficient (95% CI) | p-value |
| $\leq 140$ | 7 | 646 | 38 | 2755 | Reference | |
| 140.1 - 160 | 12 | 1327 | 19 | 2044 | 0.27 (-0.92, 1.46) | 0.66 |
| 160.1 - 180 | 13 | 1962 | 16 | 1695 | -0.18 (-1.65, 1.29) | 0.81 |
| 180.1 - 200 | 11 | 1862 | 13 | 1104 | 0.81 (-0.68, 2.31) | 0.29 |
| 200.1 + | 13 | 3042 | 18 | 1724 | 0.80 (-0.63, 2.22) | 0.28 |
| p for categorical interaction |  |  |  |  |  | 0.61 |
| Linear Weight |  |  |  |  | 0.02 (-0.01, 0.04) | 0.16 |

No. = number
\*Adjusted for gender (men were the reference group), height residuals, age (20-39, 40-59, $> 60$ ), Race (other Hispanic and other race were excluded due to small sample size), Education (Greater than, less than, or at High school level), Socioeconomic status, Comorbidities (asthma, hypertension, diabetes).
Socioeconomic status is calculated by the Center for Disease Control as a ratio of family income to the poverty line based on annual poverty guidelines specific to family size and geographic location issued by the Department of Health and Human services. Comorbidities are self-reported by patients based on if a physician has ever told them they have the condition (i.e. asthma, hypertension, diabetes).

**Table S5.**
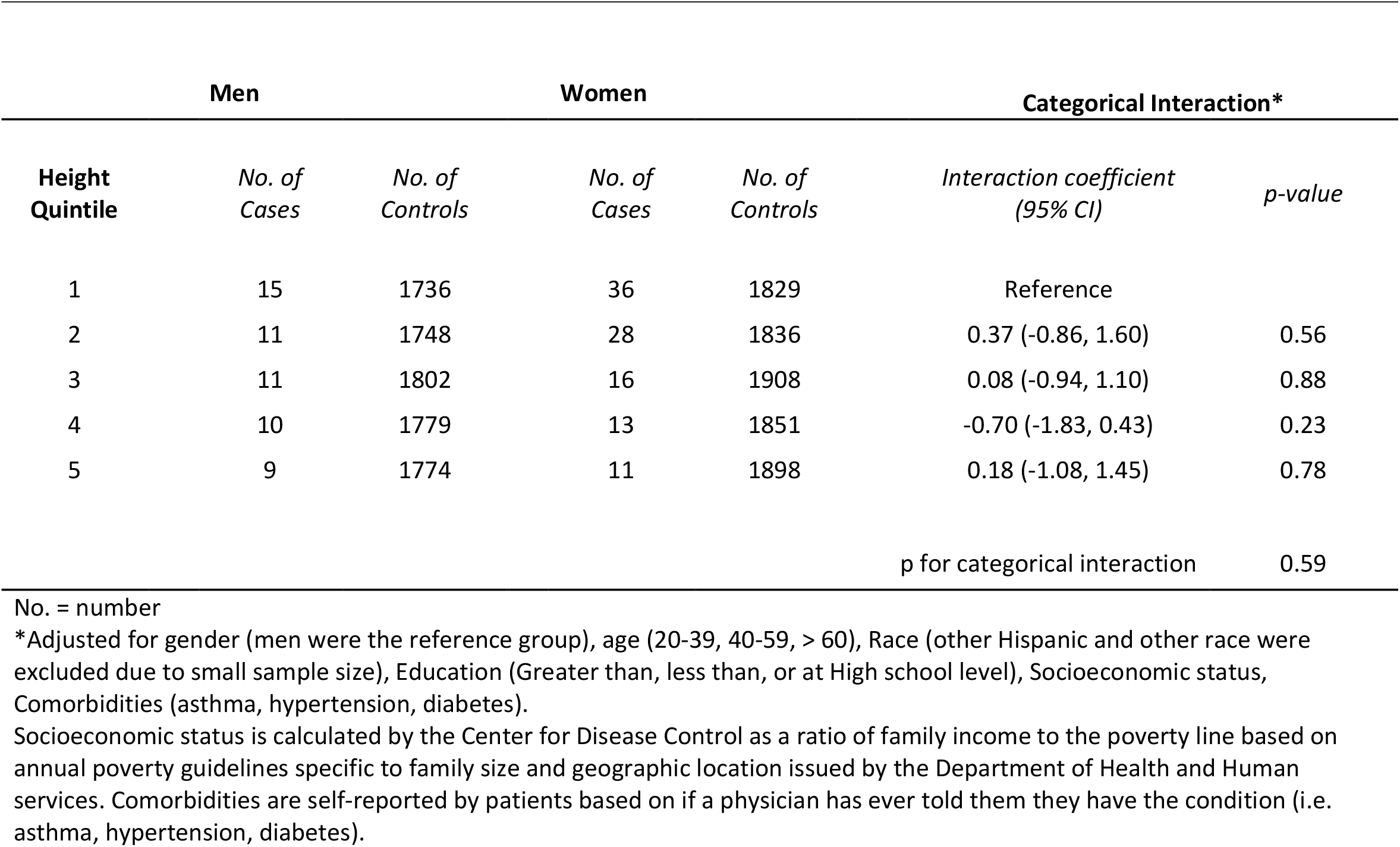
Analysis of Interaction between Gender and Gender-stratified quintiles of Height in relation to steep cornea defined by high spherical dioptric power (> 48 D) and normal level of spherical dioptric power (≤ 48 D)

## Confounding effects of anthropometric features on the association between gender and steep cornea

Prior to including anthropometric features in our model, women had greater odds of steep cornea (OR, 1.85; 95% CI 1.20-2.86; p= 0.0064). This relationship persisted with a similar degree of association upon addition of continuous BMI to the regression (OR, 1.85; 95% CI 1.20-2.86). However, when included in regression models height or weight confounded this relationship with a substantial change in the degree of association between gender and steep cornea.^12^ The degree of association between female gender and steep cornea was substantially weaker when adjusted for continuous height (OR, 0.75; 95% CI 0.43-1.30) or continuous weight (OR, 1.61; 95% CI 1.01-2.58).

## Notes

COI: Dr. Pasquale is supported by NEI EY015473 and the New York Eye and Ear Infirmary Foundation. He is a consultant to Eyenovia, Twenty-twenty and Skye Biosciences. Dr. Ahmad is supported by the New York Eye and Ear Infirmary Foundation.

### Competing Interest Statement

Dr. Pasquale is supported by NEI EY015473 and the New York Eye and Ear Infirmary Foundation. He is a consultant to Eyenovia, Twenty-twenty and Skye Biosciences. Dr. Ahmad is supported by the New York Eye and Ear Infirmary Foundation.

### Funding Statement

Dr. Ahmad is supported by the New York Eye and Ear Infirmary Foundation.

### Author Declarations

This research was deemed exempt from IRB approval by the Icahn School of Medicine at Mount Sinai IRB .

## References

1. Jinabhai A, Radhakrishnan H, O’Donnell C. Pellucid corneal marginal degeneration: A review. Contact Lens and Anterior Eye. 2011;34(2):56–63. doi:10.1016/j.clae.2010.11.007

2. Mas Tur V, MacGregor C, Jayaswal R, O’Brart D, Maycock N. A review of keratoconus: Diagnosis, pathophysiology, and genetics. Survey of Ophthalmology. 2017;62(6):770–783. doi:10.1016/j.survophthal.2017.06.009

3. François J, Goes F. Ultrasonographic study of 100 emmetropic eyes. Ophthalmologica. 1977;175(6):321–327. doi:10.1159/000308676

4. Grosvenor T, Goss DA. Role of the cornea in emmetropia and myopia. Optom Vis Sci. 1998;75(2):132–145. doi:10.1097/00006324-199802000-00017

5. Eliasi E, Bez M, Megreli J, et al. The association between keratoconus and body mass index: a population-based cross-sectional study among half a million adolescents. American Journal of Ophthalmology. Published online December 2020:S0002939420306553. doi:10.1016/j.ajo.2020.11.021

6. Jonas JB, Nangia V, Matin A, Kulkarni M, Bhojwani K. Prevalence and Associations of Keratoconus in Rural Maharashtra in Central India: The Central India Eye and Medical Study. American Journal of Ophthalmology. 2009;148(5):760–765. doi:10.1016/j.ajo.2009.06.024

7. Pan C-W, Cheng C-Y, Sabanayagam C, et al. Ethnic Variation in Central Corneal Refractive Power and Steep Cornea in Asians. Ophthalmic Epidemiology. 2014;21(2):99–105. doi:10.3109/09286586.2014.887735

8. Xu L, Wang YX, Guo Y, You QS, Jonas JB, the Beijing Eye Study Group. Prevalence and Associations of Steep Cornea/Keratoconus in Greater Beijing. The Beijing Eye Study. Yu F-S, ed. PLoS ONE. 2012;7(7):e39313. doi:10.1371/journal.pone.0039313

9. Lee KE. Association of Age, Stature, and Education With Ocular Dimensions in an Older White Population. Arch Ophthalmol. 2009;127(1):88. doi:10.1001/archophthalmol.2008.521

10. Wong TY, Foster PJ, Johnson GJ, Klein BEK, Seah SKL. The Relationship between Ocular Dimensions and Refraction with Adult Stature: The Tanjong Pagar Survey. 2001;42(6):6.

11. Engel A, Murphy RS, Maurer K, Collins E. Plan and operation of the HANES I augmentation survey of adults 25-74 years United States, 1974-1975 - PubMed. Accessed December 19, 2020. https://pubmed-ncbi-nlm-nih-gov.eresources.mssm.edu/695333/

12. Kelly A, Winer KK, Kalkwarf H, et al. Age-Based Reference Ranges for Annual Height Velocity in US Children. The Journal of Clinical Endocrinology & Metabolism. 2014;99(6):2104–2112. doi:10.1210/jc.2013-4455

13. National Health and Nutrition Examination Survey (NHANES): Vision Procedures Manual. Accessed January 18, 2021. https://www.n.cdc.gov/nchs/data/nhanes/2001-2002/manuals

14. Hoaglin DC, Iglewicz B, Tukey JW. Performance of Some Resistant Rules for Outlier Labeling. Journal of the American Statistical Association. 1986;81(396):991–999. doi:10.2307/2289073

15. Edwards M, McGhee CN, Dean S. The genetics of keratoconus. Clinical & Experimental Ophthalmology. 2001;29(6):345–351. doi:https://doi.org/10.1046/j.1442-9071.2001.d01-16.x

16. Sharif KW, Casey TA, Coltart J. Prevalence of mitral valve prolapse in keratoconus patients. J R Soc Med. 1992;85(8):446–448.

17. Beardsley TL, Foulks GN. An Association of Keratoconus and Mitral Valve Prolapse. Ophthalmology. 1982;89(1):35–37. doi:10.1016/S0161-6420(82)34857-5

18. Visscher PM, Hill WG, Wray NR. Heritability in the genomics era — concepts and misconceptions. Nat Rev Genet. 2008;9(4):255–266. doi:10.1038/nrg2322

19. Lucas SEM, Burdon KP. Genetic and Environmental Risk Factors for Keratoconus. Annu Rev Vis Sci. 2020;6(1):25–46. doi:10.1146/annurev-vision-121219-081723

20. Cutler GB. The role of estrogen in bone growth and maturation during childhood and adolescence. The Journal of Steroid Biochemistry and Molecular Biology. 1997;61(3):141–144. doi:10.1016/S0960-0760(97)80005-2

21. Weise M, De-Levi S, Barnes KM, Gafni RI, Abad V, Baron J. Effects of estrogen on growth plate senescence and epiphyseal fusion. Proc Natl Acad Sci U S A. 2001;98(12):6871–6876. doi:10.1073/pnas.121180498

22. Schuit SCE, van Meurs JBJ, Bergink AP, et al. Height in Pre- and Postmenopausal Women Is Influenced by Estrogen Receptor α Gene Polymorphisms. The Journal of Clinical Endocrinology & Metabolism. 2004;89(1):303–309. doi:10.1210/jc.2003-031095

23. Dahlgren A, Lundmark P, Axelsson T, Lind L, Syvänen A-C. Association of the Estrogen Receptor 1 (ESR1) Gene with Body Height in Adult Males from Two Swedish Population Cohorts. Brembs B, ed. PLoS ONE. 2008;3(3):e1807. doi:10.1371/journal.pone.0001807

24. Aydin E, Demir HD, Demirturk F, Caliskan AC, Aytan H, Erkorkmaz U. Corneal topographic changes in premenopausal and postmenopausal women. BMC Ophthalmol. 2007;7(1):9. doi:10.1186/1471-2415-7-9

25. Cavdar E, Ozkaya A, Alkin Z, Ozkaya HM, Babayigit MA. Changes in tear film, corneal topography, and refractive status in premenopausal women during menstrual cycle. Contact Lens and Anterior Eye. 2014;37(3):209–212. doi:10.1016/j.clae.2013.11.005

26. Soni PS. Effects of oral contraceptive steroids on the thickness of human cornea. Am J Optom Physiol Opt. 1980;57(11):825–834. doi:10.1097/00006324-198011000-00008

27. Hashemi H, Heydarian S, Hooshmand E, et al. The Prevalence and Risk Factors for Keratoconus: A Systematic Review and Meta-Analysis. Cornea. 2020;39(2):263–270. doi:10.1097/ICO.0000000000002150

28. Pearson AR, Soneji B, Sarvananthan N, Sandford-Smith JH. Does ethnic origin influence the incidence or severity of keratoconus? Eye (Lond). 2000;14 (Pt 4):625–628. doi:10.1038/eye.2000.154

29. Wagner H, Barr JT, Zadnik K. Collaborative Longitudinal Evaluation of Keratoconus (CLEK) Study: Methods and findings to date. Contact Lens and Anterior Eye. 2007;30(4):223–232. doi:10.1016/j.clae.2007.03.001

30. Sharif R, Bak-Nielsen S, Hjortdal J, Karamichos D. Pathogenesis of Keratoconus: The intriguing therapeutic potential of Prolactin-inducible protein. Progress in Retinal and Eye Research. 2018;67:150–167. doi:10.1016/j.preteyeres.2018.05.002

31. Stachon T, Stachon A, Hartmann U, Seitz B, Langenbucher A, Szentmáry N. Urea, Uric Acid, Prolactin and fT4 Concentrations in Aqueous Humor of Keratoconus Patients. Current Eye Research. 2017;42(6):842–846. doi:10.1080/02713683.2016.1256413

32. Kopelman PG. Physiopathology of prolactin secretion in obesity. International Journal of Obesity. 2000;24(2):S104–S108. doi:10.1038/sj.ijo.0801291

33. Smith SR. THE ENDOCRINOLOGY OF OBESITY. Endocrinology and Metabolism Clinics of North America. 1996;25(4):921–942. doi:10.1016/S0889-8529(05)70362-5

34. National Health and Nutrition Examination Survey (NHANES): Anthropometry Procedure Manual. Accessed January 18, 2021. https://www.n.cdc.gov/nchs/nhanes/continuousnhanes/manuals.aspx?BeginYear=2007

